# Antibody responses to SG6, AgSAP, and SAMSP1 following Anopheline salivary exposure

**DOI:** 10.1101/2025.07.14.25331506

**Authors:** Robert J. Williams, Brian D. Swinehart, Selma Abouneameh, Isaack J. Rutha, Dominick C. Msolo, Brian Tarimo, Erol Fikrig, Derrick Mathias, Billy Ngasala, Yu-Min Chuang, Jessica T. Lin

## Abstract

**Background:** Current methods to determine exposure to malaria-infected mosquitoes via entomologic investigations are technically challenging and can be inaccurate in low transmission settings. Antibody responses to mosquito salivary antigens (MSA) like gSG6-P1 have been used as biomarkers of exposure to *Anopheles* mosquito bites.

**Methods:** This study investigates two novel *Anopheles* gambiae *salivary* antigens, AgSAP and SAMSP1, as potential biomarkers of vector exposure. We evaluated the humoral response to gSG6-P1, SAMSP1, and AgSAP in a murine model and in malaria-exposed individuals with submicroscopic parasitemia across different malaria endemicity areas, seasons, and infection statuses in coastal Tanzania. We also analyzed antibody kinetics following direct skin feeding assays carried out using uninfected colony-reared *An. gambiae*.

**Results:** GSG6-P1, AgSAP, and SAMSP1 levels increased in mice at eight weeks after weekly mosquito feedings. However, human gSG6-P1 and AgSAP levels were paradoxically lower four weeks after direct skin feeding assays. SAMSP1 was the only MSA that induced a significantly higher humoral response during the rainy season, suggesting that it may be a more reliable biomarker for vector exposure in regions with multiple Anopheles species.

**Conclusions:** Mosquito salivary antigens associated with *Plasmodium* infection like AgSAP and SAMSP1 show promise as biomarkers of malaria vector exposure. However, the dynamics of IgG response against AgSAP and SAMSP1 after mosquito bites requires further study.

## BACKGROUND

Sensitive and accurate tools to measure and monitor changes in malaria transmission are essential to track progress towards malaria control and elimination goals. Current methods to determine exposure to malaria-infected mosquitoes via entomologic investigations are technically challenging and can be inaccurate in low transmission settings [1,2]. Mosquito exposure is typically assessed based on estimation of the entomological inoculation rate (EIR) [3], which is calculated by multiplying the human biting rate (HBR, the number of bites per person per year) by the sporozoite index (the proportion of captured Anopheles with sporozoites present in their salivary glands). Estimation of the EIR is inherently labor and resource intensive, requiring trained collectors and specialized laboratories. Humoral responses to mosquito salivary proteins offer a way of measuring exposure to *Anopheles* mosquitoes without laborious entomologic surveillance [4].

The most widely studied immunogenic mosquito salivary antigen (MSA) is *Anopheles gambiae* salivary gland protein 6 (gSG6), which is highly conserved among *Anopheles spp*. IgG responses to gSG6-peptide 1 (gSG6-P1) have shown promise as a biomarker of exposure to mosquito vectors across a wide range of malaria endemicity [4–6]. However, the association between HBR and gSG6-P1 is weaker when discordant Anopheles species dominate (i.e. when *An. gambiae s.l.* is not the only major vector present), such as in East Africa, where *An. funestus* is an important malaria vector, in addition to *An. arabiensis*, and *An. gambiae ss* [7]. Also, its utility as a quantitative marker for individual-level exposure remains limited [4]. Finally, gSG6 is not upregulated in infectious mosquitoes, which limits its specificity to infectious mosquito bites [8,9].

To fully realize a serologic approach to measuring Anopheline exposure, it is essential to develop additional mosquito salivary antigens and gain a better understanding of their kinetics following exposure. This study investigates additional MSAs with potential to serve as biomarkers of vector exposure, including *Anopheles gambiae* sporozoite-associated protein (AgSAP) and sporozoite-associated mosquito salivary protein 1 (SAMSP1). These proteins are directly associated with *Plasmodium* sporozoites during transmission [8–10]. Compared to gSG6, AgSAP and SAMSP1 are upregulated in infectious mosquito bites [8,9]; therefore, they may provide more specificity with regard to *infective* mosquito bites. A recent study using banked sera from Senegal showed that AgSAP and SAMSP1 reactivity peak around 2-4 weeks after clinical *P. falciparum* infection (3-5 weeks after an infectious mosquito bite) [11]. Further, those with recent malaria infection (2-4 weeks post diagnosis) had higher reactivity to AgSAP than uninfected people in the transmission season, supporting its utility as a marker for exposure to infectious mosquito bites rather than general mosquito exposure. These results highlight the promise of these MSA for serosurveillance of population-level changes in *P falciparum*-infected mosquito exposure.

We investigated the humoral response to five mosquito salivary proteins in a murine model and in malaria-exposed persons with submicroscopic parasite carriage living in Tanzania. We then measured responses to gSG6-P1, SAMSP1, and AgSAP across villages of varying malaria endemicity, across different seasons, and among people with and without malaria infection. Finally, among Tanzanian participants with submicroscopic malaria who were exposed to Anopheline bites in the context of mosquito feeding assays, we analyzed gSG6-P1, SAMPSP1 and AgSAP antibody kinetics following direct skin feeding.

## METHODS

### Mouse model of *Anopheles gambiae* bites

#### Animals

*An. gambiae* (4arr strain) mosquitoes were raised at 27 °C, 80% humidity, under a 12/12-h light/dark cycle and maintained with 10% sucrose under standard laboratory conditions in the insectary at Yale University. Swiss Webster mice were purchased from Charles River Laboratories. All animal experiment protocols were approved by the Yale University Institutional Animal Care & Use Committee (Protocol Number: 2024–07941). All animal experiments followed the Guide for the Care and Use of Laboratory Animals by the National Research Council. In all experiments, mice were housed and cared for in the Association for Assessment and Accreditation of Laboratory Animal Care International (AAALAC)-accredited animal facilities in Yale University.

#### Exposure to Anopheles gambiae bites

To determine the immunogenicity of mosquito bites, naive outbred female Swiss Webster mice were exposed to uninfected *An. gambiae* mosquito bites on a weekly basis for up to eight weeks. We let 30-50 mosquitoes take blood meals from each anesthetized mouse for 30 minutes. Sera were collected via retro-orbital sampling on Day 14 and Day 60 after first mosquito bite exposure. For retro-orbital blood collection or mosquito feeding, mice were anesthetized with intraperitoneal injection of Ketamine/Xylazine (100mg/10mg per kg body weight). When experiments were finished, mice were euthanized using a CO2 chamber in a manner consistent with AVMA guidelines for euthanasia. Following death, mice were subject to cervical dislocation as a secondary means to ensure death.

#### Antigen preparation

The *AgTRIO* and *AgSAP* sequences were designed for optimal expression using baculovirus and then subcloned into pFastBac1. The expression and purification experiments were performed by GenScript USA, Inc. SG6-P1 peptide (EKVWVDRDNVYCGHLDCTRVATF) was also synthesized by Genscript USA, Inc. Mosquito gamma interferon (IFN-γ)-inducible thiol reductase (mosGILT) and SAMSP1 were expressed and purified using insect cells [9,10]. Briefly, the coding sequences without signal peptides were inserted into an insect-cell expression plasmid pMT/BiP/V5-His, *Drosophila* S2 cells were used to express the proteins, and then protein were purified using His tag by Ni-NTA resin column and filtered through a 0.22-μm-pore-size filter.

### Clinical study population

#### Study location

This study was conducted in Bagamoyo District, Tanzania, a rural area on the eastern coast approximately 40 kilometers north of Dar es Salaam. Historically, this area had high malaria transmission but has transitioned to lower transmission over the last 15 years [12–15], with the EIR (entomological inoculation rate) dropping from >200 infective bites per person per year in the early 1990s [16] to <5 in recent entomologic survey data from the last decade [7]. Malaria transmission occurs throughout the year, with peaks typically during the long (March to May) and short (October to December) rainy seasons. We conducted malaria screening among persons ≥ 6 years of age in 2018-2021, and found that *P. falciparum* prevalence was roughly 9% by RDT/microscopy and 28% by PCR (polymerase chain reaction).

#### Study participants

This study included two groups of individuals: participants with submicroscopic malaria and endemic controls. Both groups were selected from larger, pre-existing studies.

The participants with submicroscopic malaria (N=75) were enrolled as part of the TranSMIT (Transmission of Submicroscopic Malaria in Tanzania) study between April 2019 and November 2021 [17,18]. This study prospectively screened and enrolled asymptomatic individuals aged 6 years and older from four primary schools and two health centers in Bagamoyo, Tanzania. At screening, finger-prick blood was used to make thick and thin blood smears, perform a dual antigen HPR2 and pLDH RDT (rapid diagnostic test, SD Bioline), and create 2-3 dried blood spots (DBS) on Whatman 3MM filter paper for analysis by real-time polymerase chain reaction (qPCR) targeting P. falciparum 18S rRNA [18,19]. Participants who tested positive for malaria by any diagnostic were eligible for enrollment. Venous blood was collected and approximately 2mLs plasma saved from each participant. Enrolled participants reported their age, gender, village of residence, and number of malaria episodes in the past year.

Participants who screened negative by malaria RDT but positive for *P. falciparum* by qPCR, indicating carriage of submicroscopic malaria, were further invited to complete longitudinal follow-up for 4 weeks, including weekly follow-up for symptoms and repeat parasite testing at weeks 2 and 4 and upon report of any symptoms attributable to malaria (fevers, chills, headache, body aches, malaise, nausea/vomiting). This subset of participants were provided with a thermometer to measure their temperature and were promptly treated if they developed malaria symptoms, became RDT or smear-positive, or at the end of 4 weeks if they remained parasite-positive by PCR. Plasma from 75 individuals from this group with submicroscopic malaria carriage who had specimens available at enrollment (Day 0) and Day 14 and/or Day 28 were selected for serologic analysis.

The endemic control group (N=22) participated in a SARS-CoV-2 seroprevalence study conducted in Bagamoyo, Tanzania, during the dry season July-August 2020 [20]. All had been screened for malaria as part of the TranSMIT study and tested negative by qPCR roughly three months prior to sample collection. All 22 individuals had sera drawn at a single collection time point (D0).

#### Mosquito feeding procedures

Plasma was available from 37 participants with persistent submicroscopic parasitemia who underwent mosquito skin feeding assays at two and four weeks to measure their infectiousness to mosquitoes over time. These were performed as previously described [21]. Briefly, 50 female *An. gambiae* s.s. (IFAKARA strain) 4-7 day old mosquitoes were starved 4-6 hours prior to feeding, then placed in 2 cups of 25 mosquitoes each. Cups were placed on the right and left posterior calves of each participant and allowed to feed for 15 minutes. Participants were instructed to report any discomfort, to which the feeding assay would be immediately stopped. Anti-histamine cream was applied post-feeding and was also provided for home use. Skin feeding assays were performed on Day 0 (D0), Day 14 (D14), and Day 28 (D28), with plasma collected immediately prior to skin feeding.

#### Serologic assays

A standard enzyme-linked immunosorbent assay (ELISA) with mosquito saliva proteins was conducted as described previously [9,22]. Briefly, the 96-well microplates were coated with 100µl of purified protein antigens (1µg/ml) or SG6-P1 peptide (10µg/ml) overnight. After blocking with a blocking buffer PBSTA (PBS, 0.05% tween-20, and 1% bovine serum albumin), 1:100 dilution of mouse antisera or 1:50 dilution of human antisera was diluted in PBSTA, added to the wells and incubated at room temperature for 2 hours. After washing with the washing buffer (PBS, 0.1% tween-20), horseradish peroxidase-conjugated goat anti-mouse antibody or anti-human antibody (Invitrogen) with 1:2500 dilution was used to detect total mice or human IgG. Plates were read at 450nm and 570nm, and results of each well was calculated as the result at 450nm minus the result at 570nm to remove background. Average optical density (OD) was calculated by: OD_ave_= (OD_1_ + OD_2_ - OD_blank1_ - OD_blank2_)/2, where OD_1_ and OD_2_ represent the sample duplicates and OD_blank1_ and OD_blank2_ represent the blank duplicates. Plates were tested with positive controls (pooled plasma from Mali and pooled plasma from Kédougou, Senegal) and negative controls (healthy US controls).

#### Statistical analyses

Correlation between antibody responses to different mosquito salivary antigens was measured using Spearman’s rank correlation coefficient. We used the Wilcoxon Signed-Rank test to compare two repeated samples collected at different time points after mice were exposed to weekly mosquito bites. We used the Mann-Whitney U test to compare IgG responses to salivary antigens between two independent groups. In the human clinical samples, Mann-Whitney U testing was used to compare i) those with submicroscopic malaria carriage vs. endemic controls ii) those living in villages with high vs. low malaria prevalence, based on the proportion of qPCR-positive cases in each village in the broader TranSMIT study [17], and iii) those sampled in the dry vs. rain season. Seasonality was determined using the Climate Hazards Group InfraRed Precipitation with Station data (CHIRPS)[23]. We obtained data on daily average precipitation for Bagamoyo, Tanzania. We classified dates as part of the dry season if the preceding 28-day precipitation totals fell below 60mm as defined in the Köppen Climate Classification [24]. We used the Friedman test to compare repeated samples collected at different time points after human participants were exposed to direct skin feeding assays, as well as to compare subgroups within this group of participants. All p-values are representative of a two-tailed test. We performed all analyses using GraphPad Prism 9, R version 4.4.1, and Microsoft Excel.

## RESULTS

### Humoral responses in mice against mosquito salivary proteins after exposure to mosquito bites

Since mosquitoes inject saliva proteins when probing, mosquito-exposed mammals can potentially generate humoral responses against different salivary proteins after repeated mosquito bites. In 13 mice exposed to 30-50 female uninfected *An. gambiae* mosquito bites on a weekly basis, increased IgG reactivity against SAMSP1 and AgTRIO was noted at two weeks, with a trend towards increased IgG reactivity to mosGILT, but not against SG6-P1 or AgSAP at this early time point. However, after eight weeks of weekly mosquito bite exposure, elevation of IgG against all salivary proteins was noted in the exposed mice, except for AgTRIO, for which a trend towards increased reactivity remained (**Figure 1**).

**Figure 1.**
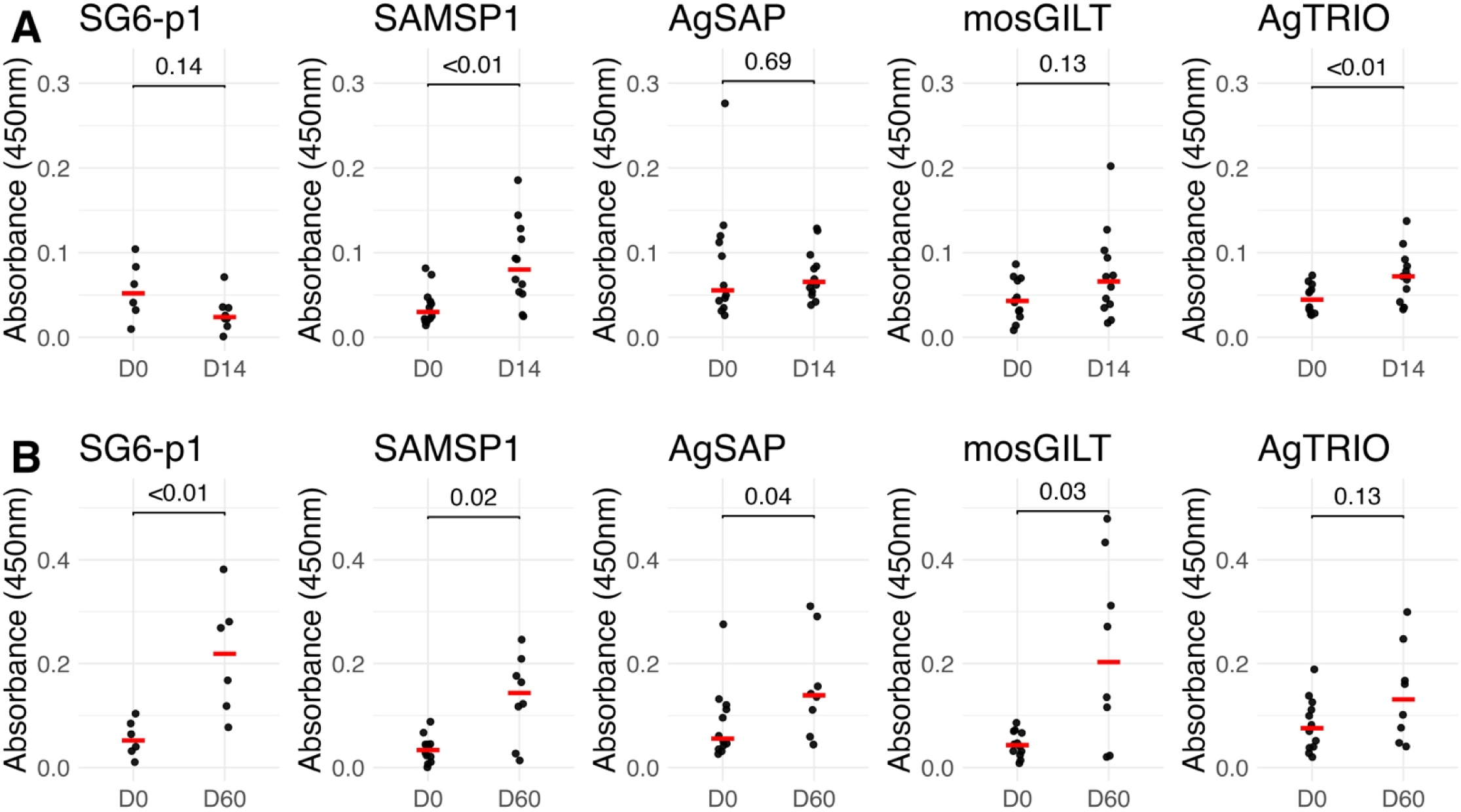
IgG response to salivary proteins in Anopheles-bitten mice. Swiss Webster mice were exposed to 30-50 mosquitoes bites weekly and IgG responses against mosquito salivary antigens were measure by ELISA. **(A)** IgG responses at D0 and D14 (two mosquito feeds) and **(B)** IgG responses at D0 and D60 (eight mosquito feeds).

### Association of human anti-salivary protein IgG responses with malaria and transmission season

Human IgG responses to the same five Anopheline salivary antigens were measured among 97 residents of rural coastal Tanzania greater than 5 years of age, including 75 asymptomatic individuals with submicroscopic malaria carriage identified by *P. falciparum* qPCR and 22 endemic controls recruited during the dry season who had recently screened PCR-negative for malaria. In this group, IgG responses to AgSAP and SAMPS1 were correlated with responses to AgTRIO (r = 0.60) and mosGILT (r=0.80), respectively (Figure S1). Thus, subsequent experiments focused on SG6-p1, AgSAP, and SAMPS1 (distribution of responses depicted in Figure S2).

Compared to the endemic controls, those with evidence of submicroscopic malaria carriage appeared to display higher anti-salivary responses to all three antigens, though these trends did not reach significance (**Figure 2A**). Anti-salivary responses to SG6-p1 and AgSAP were also higher in individuals living in villages with high malaria prevalence (N=33) compared to individuals living in villages with lower malaria prevalence (N=26) (**Figure 2B**).

**Figure 2.**
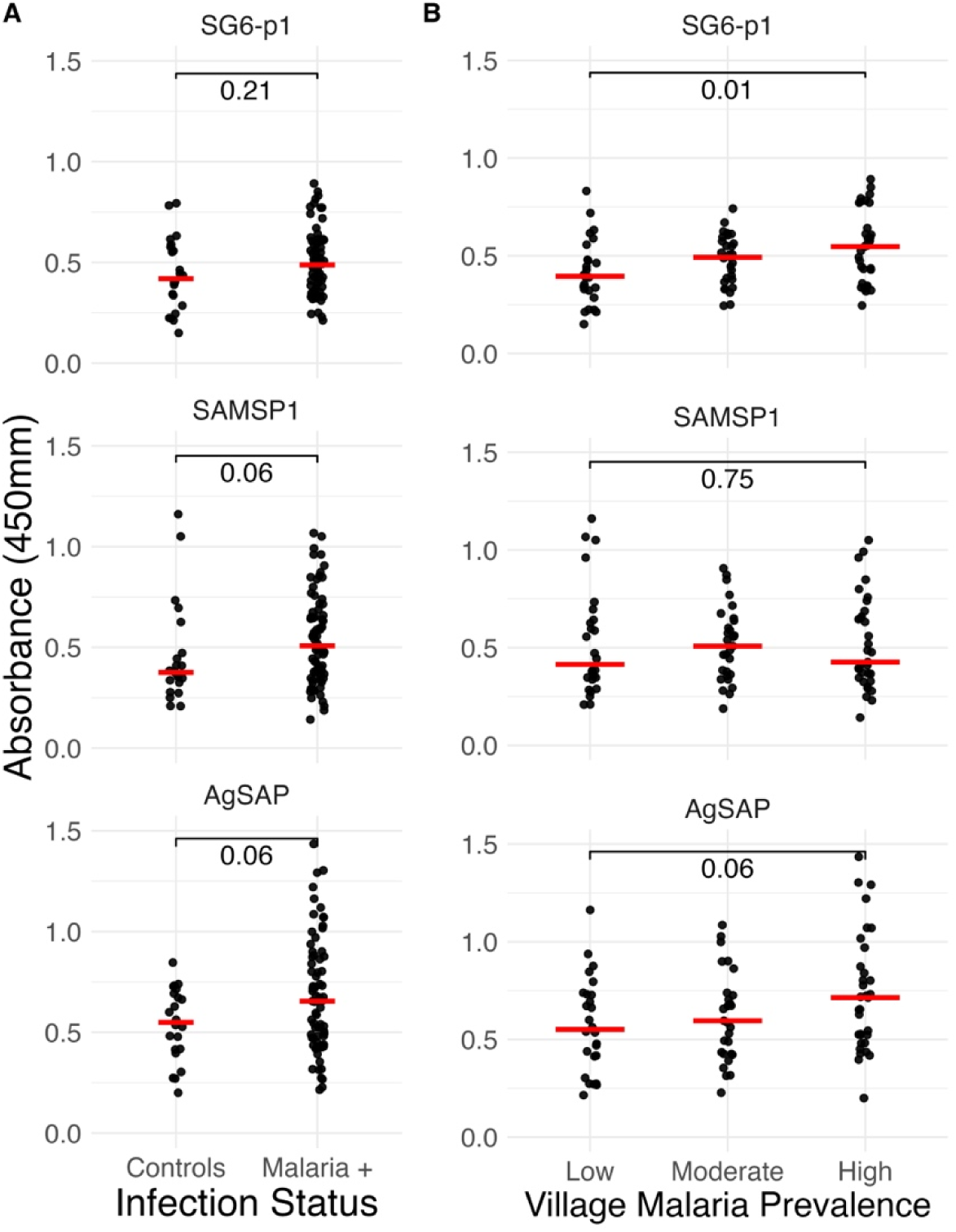
Association of anti-salivary responses with malaria exposure. (A) IgG responses of participants with submicroscopic malaria carriage (N=75) compared to endemic controls (N=22). (B) IgG responses of by malaria village prevalence. Malaria prevalence was categorized as Low (19-23% qPCR positive), Moderate (24-28%), and High (29-45%) based upon qPCR prevalence from 2018-2021 as part of the Project TranSMIT study.

Compared to antibody responses during the dry season, participants enrolled during the rainy season demonstrated increased IgG reactivity to SAMSP1 and potentially AgSAP, but no seasonal association was found for SG6-p1. (**Figure 3**). These differences were greater in residents living in high malaria prevalence villages (Figure S3). There was no association between anti-salivary responses and sex or age in the cohort (Figure S4).

**Figure 3.**
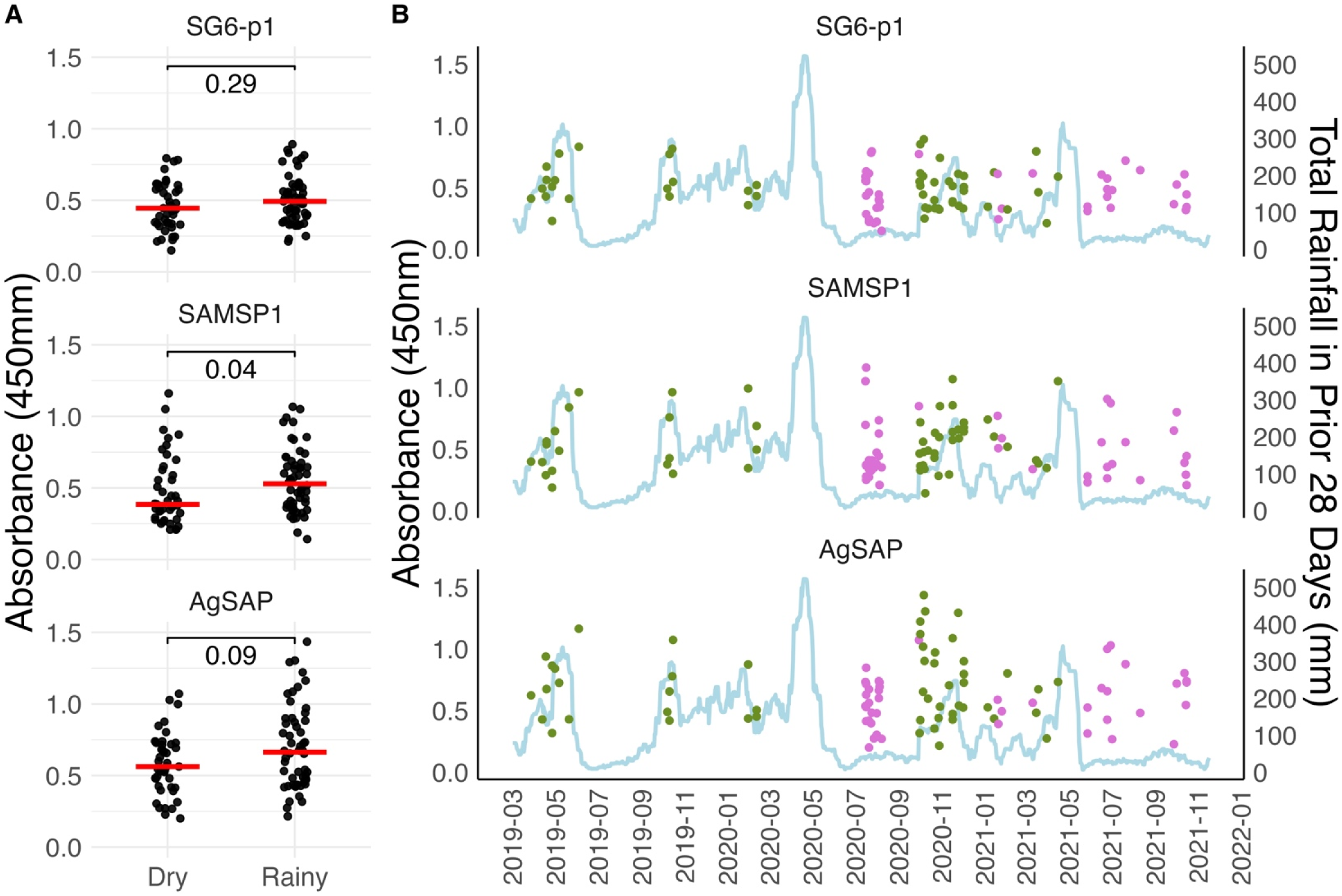
Association of anti-salivary response and enrollment season. **(A)** IgG responses by dry and rainy season. The red horizontal line represents the median for each group. The rainy season was defined as a month period with > 60mm of rainfall. **(B)** IgG responses by date and season (dry in purple, rainy in green). The light blue line represents the total rainfall in the prior 28 days.

### Human anti-salivary IgG responses following mosquito direct skin feeding

As part of the TranSMIT study [17,18], 37 participants with submicroscopic malaria were exposed to Anopheline saliva through direct skin feeding assays with 50 female *Anopheles gambiae* fed on the posterior calves for 15 minutes. After two direct skin feeding assays spaced two weeks apart (one at enrollment day 0, and another 2 weeks later at day 14), the median anti-SG6 and anti-AgSAP antibody response at day 28 paradoxically declined (**Figure 4**). This decline was most prominent in those living in villages with high malaria prevalence (n=13), those who were sampled during the rainy season (n=22), females (n=24), and those who reported having one or more malaria episodes in the past year (n=24) (Figure S5).

**Figure 4.**
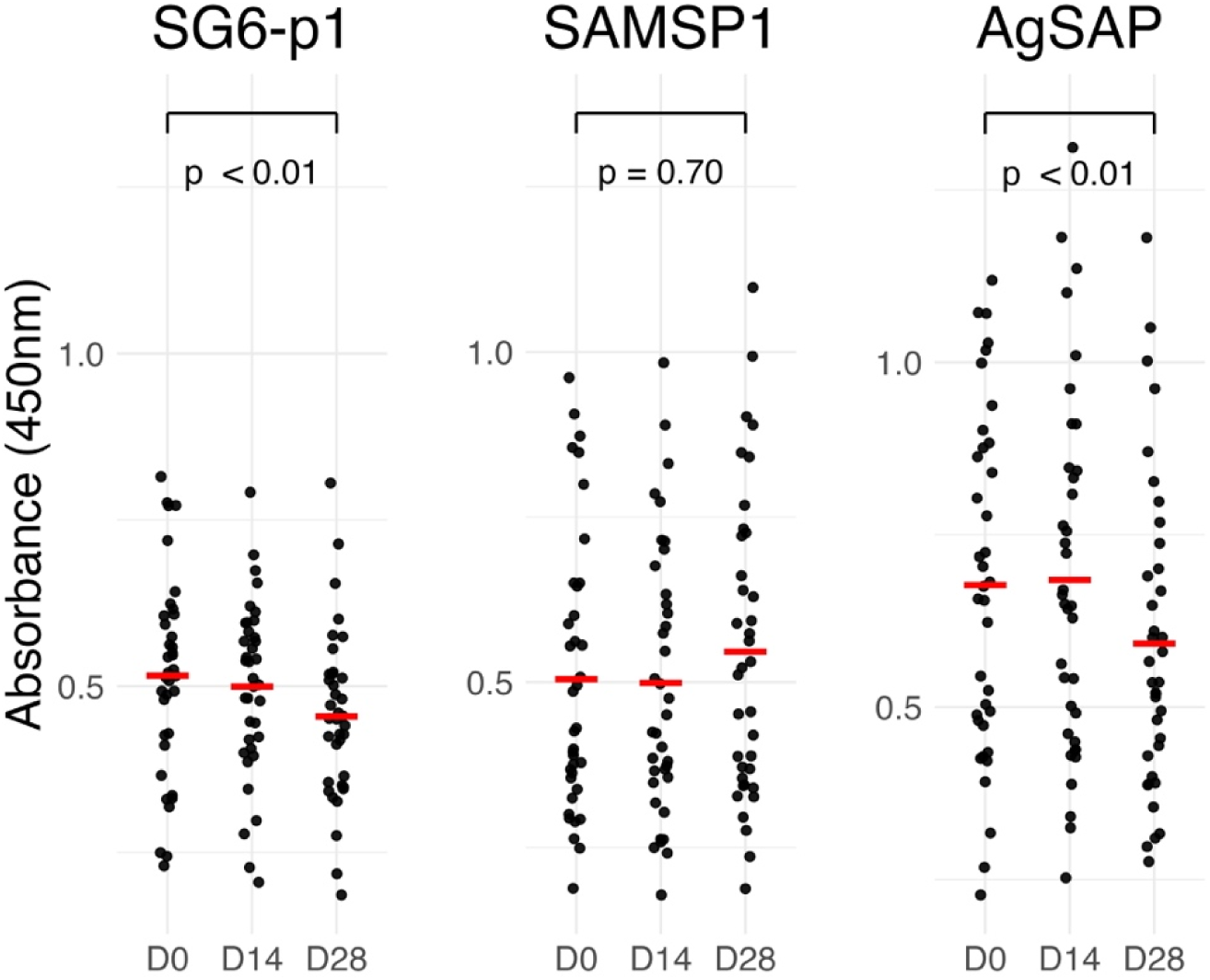
Anti-salivary IgG responses in those with submicroscopic malaria carriage after direct mosquito skin feedings assays at enrollment (D0) and 14 days later (D14).

On an individual basis, the patterns observed from two feeding assays conducted two weeks apart were quite heterogeneous. In terms of the most common patterns for each antigen, anti-SG6p1 showed a sequential decline in roughly half of skin-fed participants (18/37), while anti-SAMSP1 exhibited a sequential increase in 30% (11/37), and AgSAP showed boosting at two weeks, followed by a decline at four weeks in 43% (16/37) of participants (**Figure 5**). We could not discern that individual patterns of boost/decline were associated with participant characteristics. However, 12/37 (32%) participants exhibited the same pattern for all three antigens (Figure S6).

**Figure 5.**
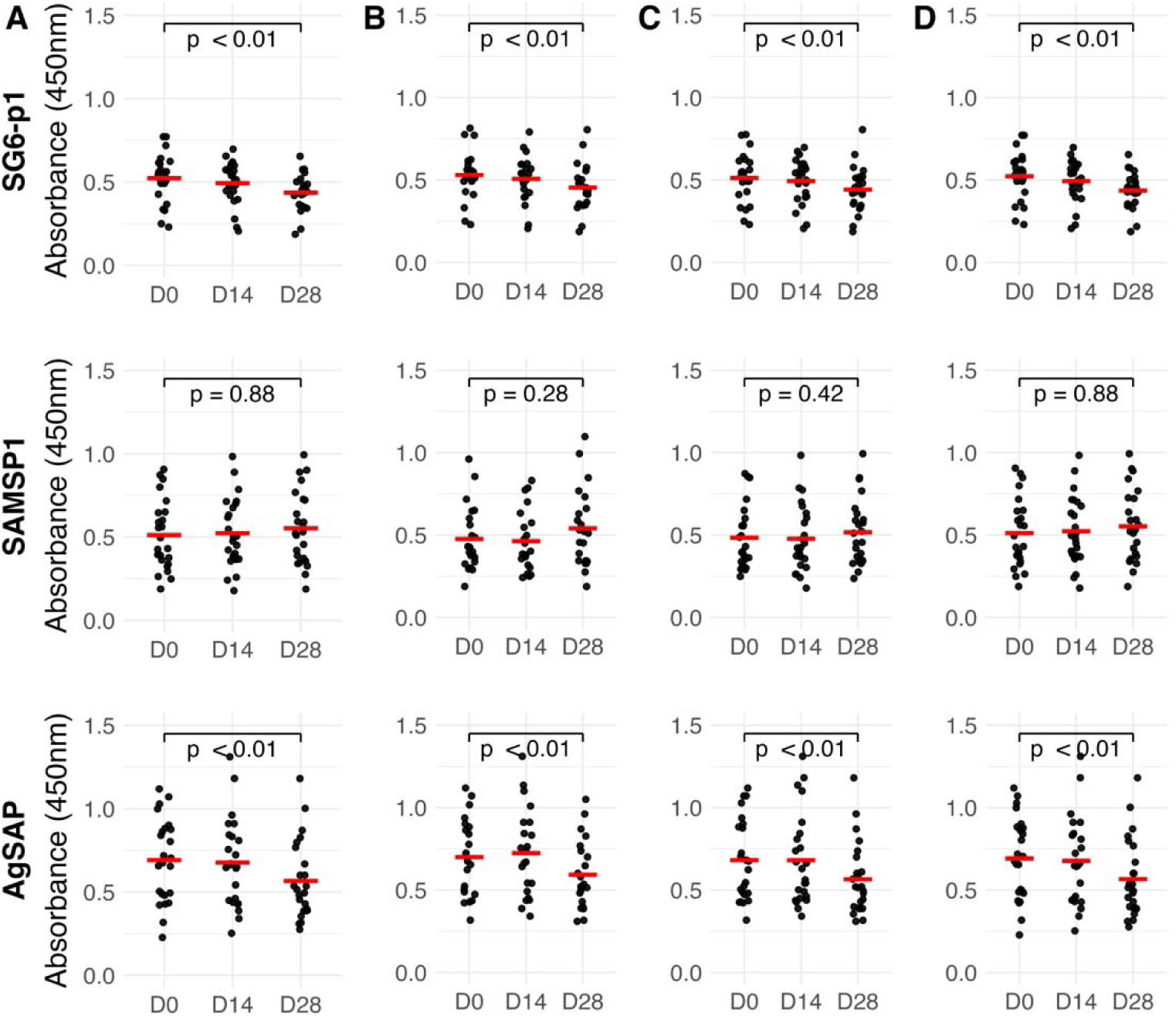
Individual anti-salivary IgG responses at Day 14 and Day 28 after mosquito skin feeding assays at Day 0 and Day 14. Longitudinal IgG levels are shown for each participant, with the predominant pattern for each mosquito salivary antigen depicted in black, whether sequential decrease (SG6), sequential increase (SAMSP1), or “boost” at two weeks then subsequent drop at four weeks (AgSAP).

## DISCUSSION

Mosquito salivary antigens (MSAs) have shown promise as indicators of human exposure to mosquito vectors [4]. Their measurement could help assess the efficacy of vector intervention strategies, particularly in natural settings where multiple factors may influence malaria transmission, or when multimodal interventions are being implemented and tested. In this study, we aimed to evaluate whether *An. gambiae* MSAs elicit humoral responses and whether these responses are related to the level of human-vector exposure, focusing on two novel markers, AgSAP and SAMSP1, in addition to a well-studied marker, gSG6-P1.

Using a murine model, we demonstrated that SAMSP1 and AgSAP are antigenic after repeated exposure to *An. gambiae* bites over an 8-week period, with anti-SAMSP1 IgG rising as soon as two weeks following mosquito exposure. A recent study from Senegal [11] did not find any differences between AgSAP and SAMSP1 responses in areas of low versus moderate malaria transmission. When comparing antibody responses in those living in high vs low malaria prevalence villages, we did not find differences in SAMSP1 levels, but did find increased reactivity for both AgSAP and gSG6-P1 in the higher prevalence villages, though this finding was borderline significant for AgSAP (p=0.06). Our findings are consistent with several studies that have reported higher gSG6-P1 levels in areas of higher malaria prevalence [25–27].

While our study lacks data on entomological exposure, several studies have shown an increase in gSG6-P1 levels during the rainy season, when HBR is higher [4,27–29]. In contrast, we did not find an association between gSG6-P1 and seasonality. This discrepancy may be due to the multiple *Anopheles* vectors that exist in coastal Tanzania, as the association between HBR and gSG6-P1 levels is strongest when *An. gambiae s.l.* is the only dominant vector species [4]. Alternatively, we found that SAMSP1 levels were significantly higher during the rainy season, suggesting that SAMSP1 may be a more reliable biomarker for vector exposure in regions with multiple Anopheles species. The *An. gambiae* SAMSP1 gene shares slightly greater sequence identity (82%) with its *An. funestus* homologue than An. gambaie SG6 (77%) or AgSAP (53%) for their *An. Funestus* homologs, and even shares 64% sequence identity with its homolog in *Aedes aegypti* [11].

GSG6-P1, AgSAP, and SAMSP1 levels were all higher in individuals with submicroscopic parasitemia compared to endemic controls, though none of these differences reached statistical significance (p=0.21, p=0.06, p=0.06, respectively). Several studies have shown increased gSG6-P1 levels in malaria infected individuals when compared to uninfected individuals [26,30–33]. AgSAP levels have been found to be higher in individuals with recent malaria infection (2-4 weeks after clinical malaria infection) in low and moderate transmission areas, whereas SAMSP1 was found to only be higher in recent infection in areas of low malaria transmission [11]. AgSAP and SAMSP1 are upregulated in infectious mosquitoes and a bite from an infectious mosquito could induce a greater immune response than a bite from an uninfected mosquito [9].

We were surprised by the decline in gSG6-P1 and AgSAP levels after direct skin feeding assays. The Senegal study showed that AgSAP and SAMSP1 levels are highest 2-4 weeks after clinical malaria and decrease by 3 months after infection [8,11]. As all of our participants who participated in the skin feeding assays had asymptomatic submicroscopic parasitemia, we do not know when they were infected, and it is possible that we are simply observing the gradual decline in these antibody levels from their peak levels. This may be supported by our finding that those expected to have greater malaria exposure (those living in high prevalence malaria villages sampled in the rainy season with a history of malaria) were the most likely to have declining IgG levels. Alternatively, uninfected mosquitoes may not induce a strong humoral response; levels may only increase as a result of exposure to *infected* mosquitoes. It is also possible that the stimulus of 50 mosquito bites was not large enough to induce a measurable response. Regardless, most studies that have reported a relationship between MSA levels and malaria risk are descriptive [4], and our findings highlight the need for an increased understanding of mosquito salivary antibody kinetics [34].

Our study has several limitations. A primary limitation is the lack of data on entomological exposure. We inferred exposure based on malaria prevalence in each village, which may not accurately represent the true human-vector exposure relationship. Additionally, the absence of a control group in the direct skin feeding assay is a significant limitation. It is possible that uninfected mosquitoes do not elicit a strong immune response to AgSAP and SAMSP1. Without a control group, we cannot determine if the observed decrease in antibody levels is expected in cases of submicroscopic parasitemia. The study would also have benefitted from samples collected at later points after mosquito feeding, since not enough time may have elapsed at four weeks to evaluate responses induced by the second mosquito feeding at two weeks.

## CONCLUSIONS

Malaria is a major health challenge in sub-Saharan Africa. Accurately estimating the risk of malaria transmission is essential for deploying and monitoring effective control, management, and elimination strategies. This study demonstrates that AgSAP is a promising biomarker for exposure to anopheline bites and could be a valuable addition to serological tools for estimating human-vector contact.

## Data Availability

The datasets used and/or analyzed during the current study are available from the corresponding author upon request.

## DECLARATIONS

## Acknowledgements

We thank the study participants as well as the staff at the schools and health centers in Bagamoyo for their support. We thank the study teams at Muhimbili University of Health and Allied Sciences (MUHAS) for coordinating the field activities and the Ifakara Health Institute for coordinating the mosquito experimental infections.

## Authors’ contributions

JL and YC conceptualized and designed the study. YC led the experimental mouse work, including acquisition, analysis, and interpretation of data. BN led the field team for subject enrollment, including acquisition of data. DM, BT, and IR carried out the mosquito feeding assays, including acquisition of data. SA and BS carried out the molecular work, including acquisition and interpretation of data. BS and RW carried out data analysis. RW, JL, YC, and BS drafted the manuscript. DM, SA, and EF helped revise the manuscript. All authors read and approved the final manuscript.

## Ethics approval and consent to participate

This study was approved by institutional review boards at the University of North Carolina (ID 276606), Tanzania National Institute for Medical Research (NIMR/HQ/R.8a/Vol.IX/3150), Ifakara Health Institute (IHI/IRB/33–2018), and Muhimbili University of Health and Allied Sciences (MUHAS/DA.282/298/01/C). Informed consent in Kiswahili was obtained prior to screening for malaria and for enrollment into the mosquito feeding portion of the study.

## Consent for publication

Not applicable

