## Supplementary figures and images for "Antibody responses to SG6, AgSAP, and SAMSP1 following Anopheline salivary exposure"

### Supplement

#### **SUPPLEMENTARY MATERIAL**

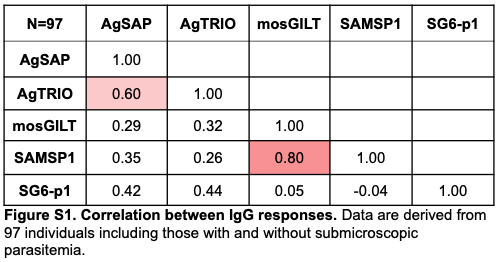

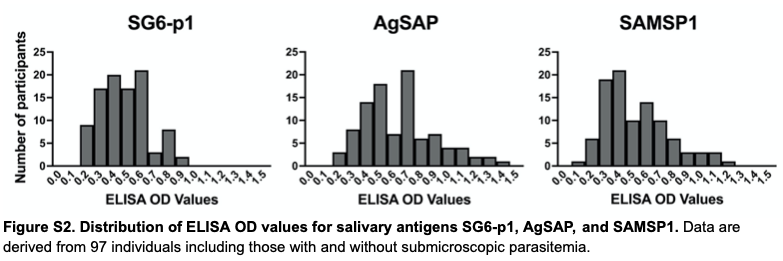

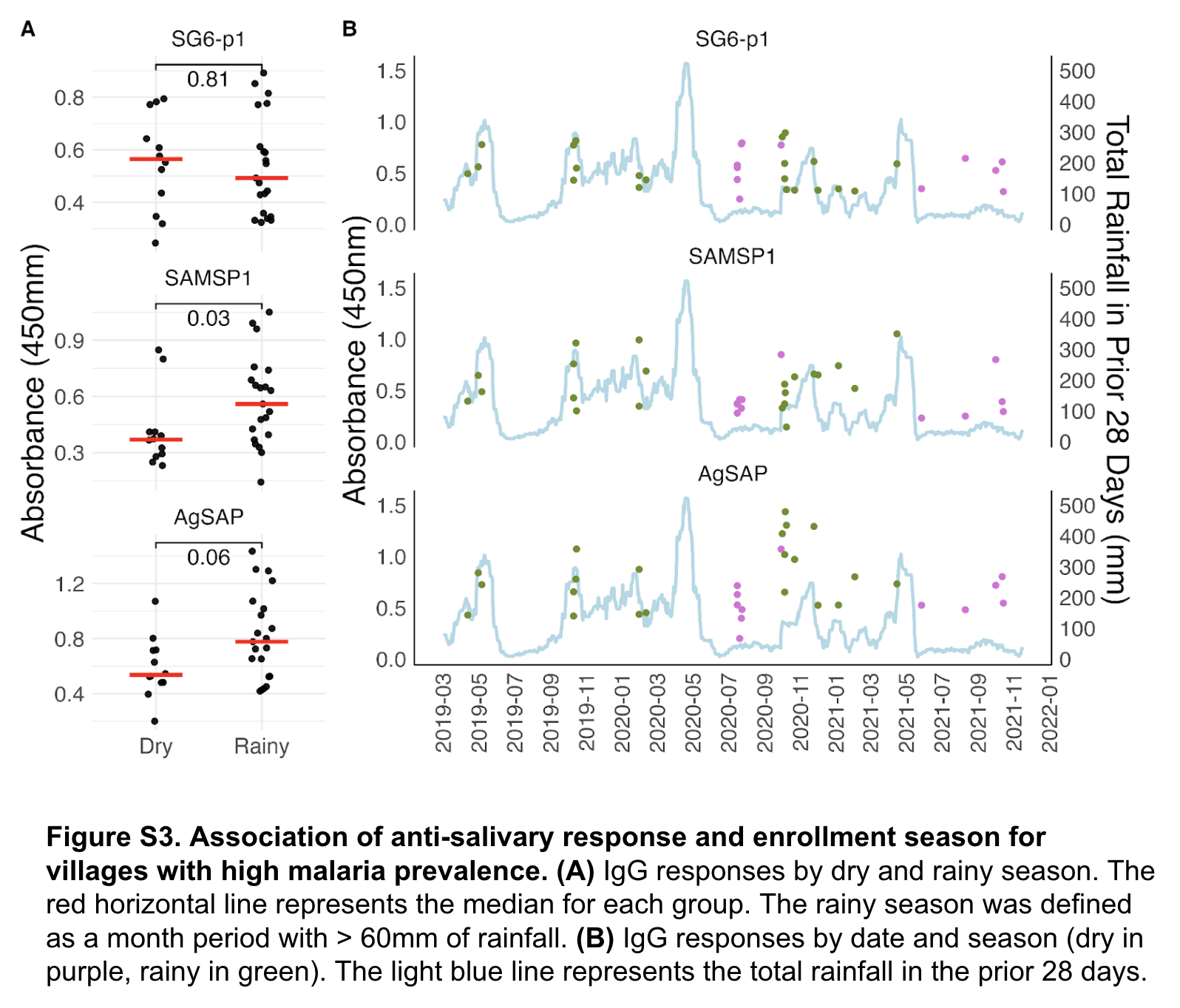

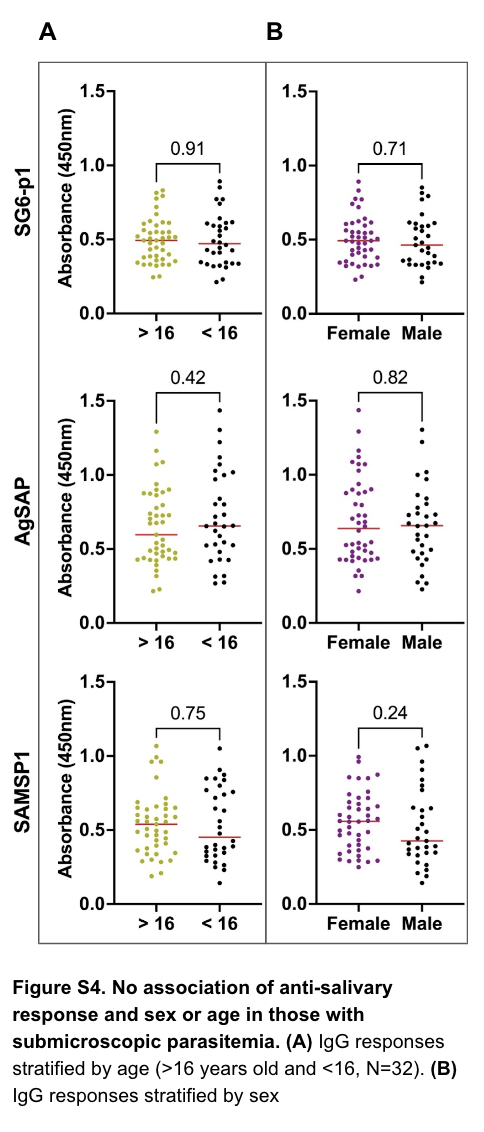

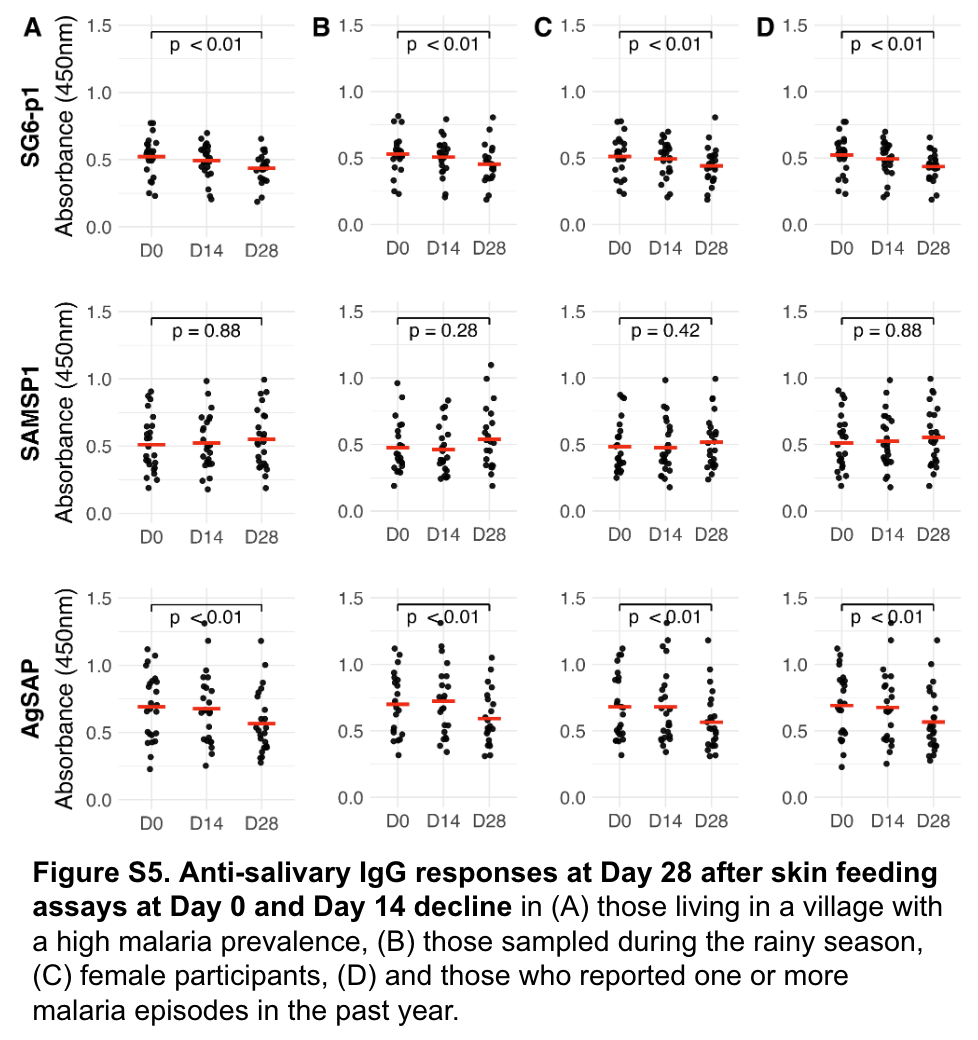

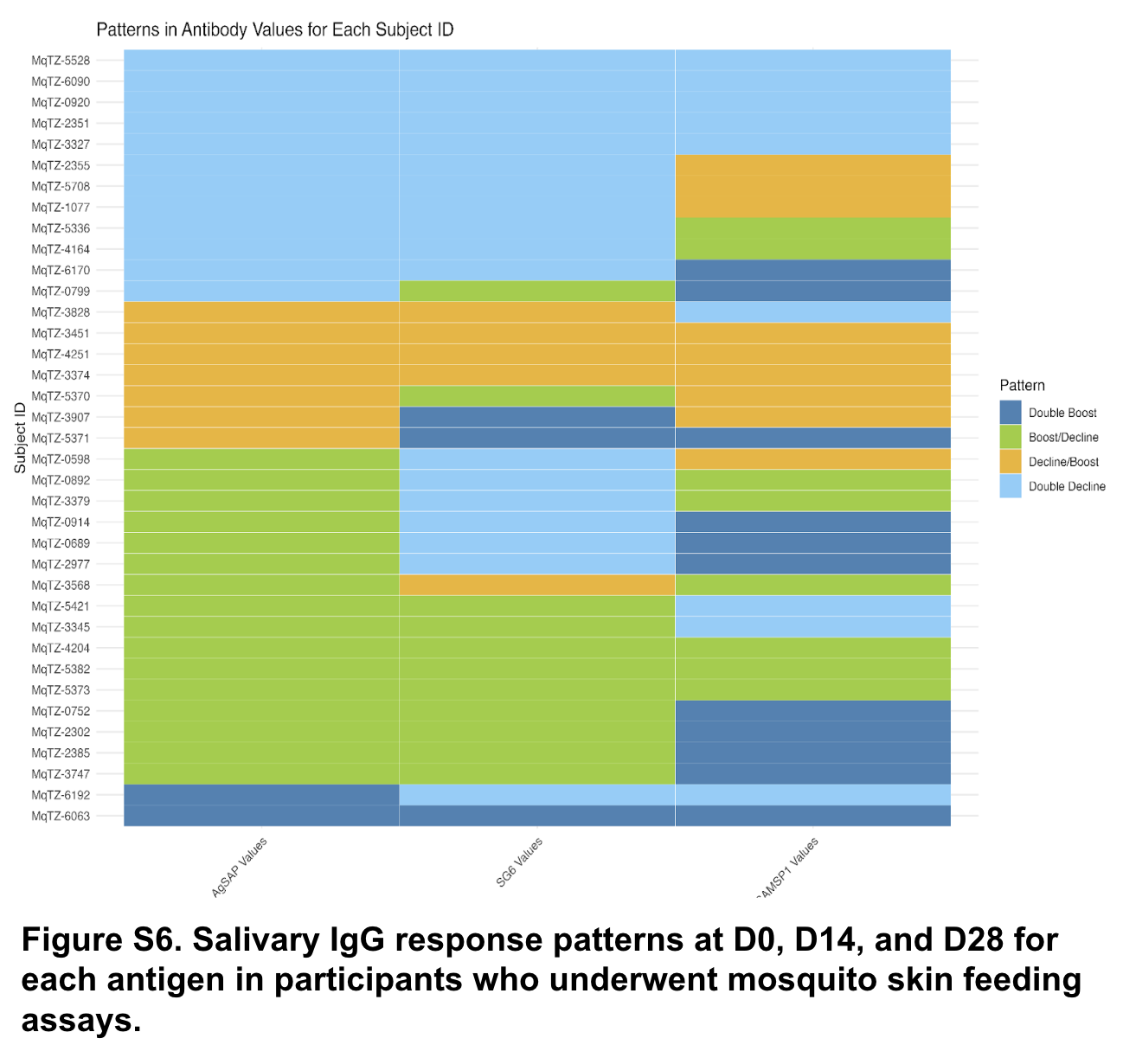
